# Mortality rates in matched cohort, pseudo-randomised and randomised trials of convalescent plasma given to COVID-19 patients

**DOI:** 10.1101/2020.11.19.20234757

**Authors:** Amar Ahmad, Marwa Salsabil, Tim Oliver

## Abstract

**Introduction:** For more than 80 years convalescent or immune sera has been used in severe life threatening infections. Since March of this year a rapidly increasing number of publications have reported series of Convalescent plasma (CP) investigations in severely ill COVID-19 patients.

**Objective:** a brief CP scoping review focusing on early mortality

**Methods:** We searched available data bases. Three randomised trials, two pseudo-randomised observations and twelve matched cohort studies were identified. Random-effects meta-analyses were performed on extracted data

**Results:** A total of 2,378 CP treated and 5188 “controls” in 17 studies. Individually only two studies were significant for reduction of deaths to 30 days, but all showed a similar percentage reduction. When pooled, meta-analysis was undertaken. It showed that the overall reduction of death was significant for all series RR 0.710 (p=0.00001), all matched cohort series RR = 0.610 (p-value = 0.001) and the two pseudo-randomised series RR 0.747 (p=0.005) but not the three technically inadequate randomised trials, RR 0.825 (p=0.397). In two of these randomised trials, there was faster clearance of Viral DNA at 72 hours after CP than placebo

**Conclusion:** It is hoped the significance of this less than perfect data will increase interest in completing the delayed randomised trials as the results suggest they could be better than currently licenced drugs. Given increasing published evidence of increased risk of both diagnosis and death from COVID-19 in patients with severe Vitamin-D deficiency, future studies should also study influence of Vitamin-D status of donor and recipient on outcome.

| What is known about the Convalescent Plasma (CP)? | What is new? | What are the key questions for future work on the topic? |
| --- | --- | --- |
| Since the 1920s cohort studies have suggested that CP is an effective treatment in epidemic lethal infectious diseases but the high mortality had hindered completion of randomised trials | As only 13.8% of the COVID-19 cases reviewed were in randomised trials this paper has explored the role of pseudo-randomisation (as a form of non-genetic mendelian randomisation) and RR instead of OR to give earlier indication of the case numbers needed for randomised trials | Increased publicity about the need to complete recruitment to randomised trials, and if positive immediate vs deferred use trials to clarify how urgently CP needs to be given. Investigate the role of treating Vitamin D deficiency in recipient and donor and the use of vaccines to improve the titre of low antibody titre donors to increase the pool of available donors and development of anti- SARS-CoV-2 monoclonal antibodies |
| There are only 2 drugs licenced specifically for use in COVID-19 illnesses, one more suited to early COVID-19 and one in more advanced cases. In the trials analysed there are very different mixtures of licenced and non-licenced drugs producing considerable risk of biases. | This review is the first to compare the results after use of CP with the reported trial data on Dexamethasone and Remdesivir and draw attention to CPs apparent faster action in clearing evidence of SARS-CoV-2 than Remdesivir | Under an individual patient data systematic review in order to look at individual drug interactions with CP action |

## Introduction

Since the first reports of a cure for terminal children with Diphtheria induced laryngeal obstruction with immune serum in 1898 (1) and the successes in 1917-18 flu epidemic with use of convalescent serum (2), this approach has been successfully used in bacterial diseases such as meningitis (3) and viral epidemics (4) most recently including Ebola (5) Unfortunately to date, most of the success has depended on case-control studies (6) rather than randomised trials to properly establish if there is a real benefit from this approach. The first report of its potential benefit compared to matched untreated patients in severe COVID-19 infection was in April 2020 (7). Since then there has been increasing interest (8, 9) and three ongoing Cochrane reviews which have demonstrated that un-matched cohort studies are still the most frequent reports (10-14). The most recent Cochrane report identified 2 underpowered randomised control trials and 8 studies with matched controls, though because of identified bias being high in the matched studies they only assessed mortality in the randomised trials, which though reduced was not significant due to lack of numbers.

### Objectives

Discovering an increasing number of articles since 19^th^ August the search date in the recent Cochrane study, led to a literature search and review that only focused on the cumulated mortality data from these reports to attempt to compare them with those in randomised and “pseudorandomised” studies and then compare the results with those from drugs currently licenced for COVID-19 in the hope of increasing interest in completing outstanding randomised trials during the winter surge in COVID-19 infection that has now taken off.

## Methods

Searches of Pubmed, EMBASE, Web of Science and ClinicalTrials.gov have been undertaken using the following terms: Convalescent Plasma and COVID-19 and year published 2020. At the last search 22.10.2020, 153 references were identified through Medline 143 through EMBASE, 363 through Web of Science and 96 from ClinicalTrials.gov currently recruiting and 16 completed. To be included the title or abstract had to indicate that their data related to a randomised, pseudorandomised (see below for discussion of rationale for inclusion) or Matched Cohort study. To compared the results with data on established COVID-19 therapies, three randomised trials with Remdesivir (15-17) and one trial with Dexamethasone (18) were reviewed

From the review, a total of 16 papers with identified original data comparing convalescent plasma and randomised (n=3), pseudo-randomised (n=2) or matched cohort controls (n=11) were read as full papers. A seventeenth paper (19) was identified through the review of Joyner (8). The original article gave no indication of number of controls (they did indicate in the manuscript that the results compared favourably with their institutional database). For this paper, as the review by Joyner documented such data but without information on follow up, this information has been used in this review.

The decision for exploring the data from pseudo-randomised series was because If a treatment is not randomised but is given “in a perceived” uniform cohort manner but post-treatment completion, a significant prognostic factor unsuspected at the trial’s initiation emerges as a risk factor that influencing survival, this factor could not be deemed to have influenced actual allocation. In such a case, Joyner et al (https://pubmed.ncbi.nlm.nih.gov/32817978/) considered those allocated by chance to the extremes of the perceived specific new risk factor, are considered to be pseudo-randomised and therefore of possibly of more value for examining this role of this specific factor than a conventional matched cohort study though still not as good as specific randomisation.

If two such significant prognostic factors unsuspected at the trial’s initiation emerges as separate risk factors that influencing survival and prove synergistic in their interaction, could the outcome of double positives versus double negatives in such a “Pseudo-pseudo” Randomisation of two prognostic factors provide a closer approximation to true randomisation than Pseudo-randomisation? In using two prognostic factors it is following the concept of Mendelian randomisation except it uses prognostic factors instead of gene definitions. Of course, it still does not reach the reliability of a randomised controlled trial, but the composite control double negatives could be closer to a true placebo effect than a randomised trial without a precise placebo because of no way of determining the impact of the placebo effect.

In the largest “pseudo-randomised” study (20), those receiving the lowest and highest titre plasma were compared. In the smaller study (21), the comparison was an arbitrary cut-point based on the levels of serum IgG against the spike protein of SARS-Cov-2 measured by an enzyme-linked immunosorbent assay (ELISA) using a validated commercial kit (Euroimmun AG, Luebeck, Germany, product number EI 2606-9601G) comparing the upper 39% versus the lower 61%. Two Chinese studies (22, 23) were pooled as the first had pooled small numbers of three early reports with 19 patients with 10 matched controls and the latter reported on the formal phase II study which followed.

For each study, the proportion that died and follow up after treatment and control groups were tabulated and absolute and relative risk calculated. The same information was obtained from three Remdesivir and one Dexamethasone trials all randomised to compare with the outcome from convalescent plasma data.

### Statistical Methods

Random-effects meta-analyses were performed to estimate combined average effect of deaths after COVID-19 convalescent plasma. The restricted maximum-likelihood (REML) estimator was used by means of the rma R command (24). The percentages of deaths after COVID-19 convalescent plasma were compared to those in matched controls. The main statistical analysis included eleven matched cohort comparisons. The combined relative risk (RR), which is the ratio of the percentage of death in the plasma group to the percentage of death in the standard group were computed (25). A secondary analysis was carried out by adding three randomised and two pseudo-randomised studies to the ten matched cohort studies. Sub-group meta-analyses were carried out on the randomised and pseudo-randomised set respectively. A RR statistic was used for all meta-analyses (26). All meta-analyses estimations were weighted with inverse-inverse variants (R-rma default). The corresponding z-statistic, which is the estimated effect divided by its standard error, and its p-value were computed. Between study, heterogeneity was assessed by the Q-statistic test and I^2^ statistic which is the ratio of the total heterogeneity divided by the total variability (27).

A mixed-effects model (meta-regression) was performed with the REML estimator including 13 studies and a categorical moderator was used to distinguish the three sets (i.e. the nine matched cohort studies, two pseudo-randomised studies and the two randomised trials) (28)

All applied statistical tests were two-sided, p < 0.05 were considered statistically significant. No adjustment for multiple comparisons was made. Statistical analyses were performed in R version 4.0.2 (29).

### Ethics

With all the data analysed already in the public domain and no need to involve actual patients in the analysis, it was felt unnecessary to seek ethical approval for the study

## Results

A total of 2,378 Convalescent plasma (CP) treated and 5188 “controls” were identified in the 17 studies and for the analysis, the studies of Chan et al (30) and Xie et al (23) were merged making 16 comparisons (Table 1). This table also indicated number and percentage of deaths and the duration of follow up which varied from >5 to 30 days (median 17.5 days). Absolute reductions were calculated from the difference between percentage deaths in Controls vs CP groups and ranged from 2.4% to 23.9% (median 11.5%) for the 11 matched cohort comparisons while it was –0.9%, 8.3% and 11.6% for the randomised trials and 7.3% and 12.8% for the pseudorandomised studies. The overall absolute risk reduction was median 10.8%, mean 11.72%. Using the difference between the median frequency of deaths in the treated (13.4%) versus controls (26.7%), the sample size of a randomises trial needed to treat to prove this difference would be 141 in each arm while in the worst-case scenario 1.9% versus 4.3% it would be 817 per arm.

**Table 1.**
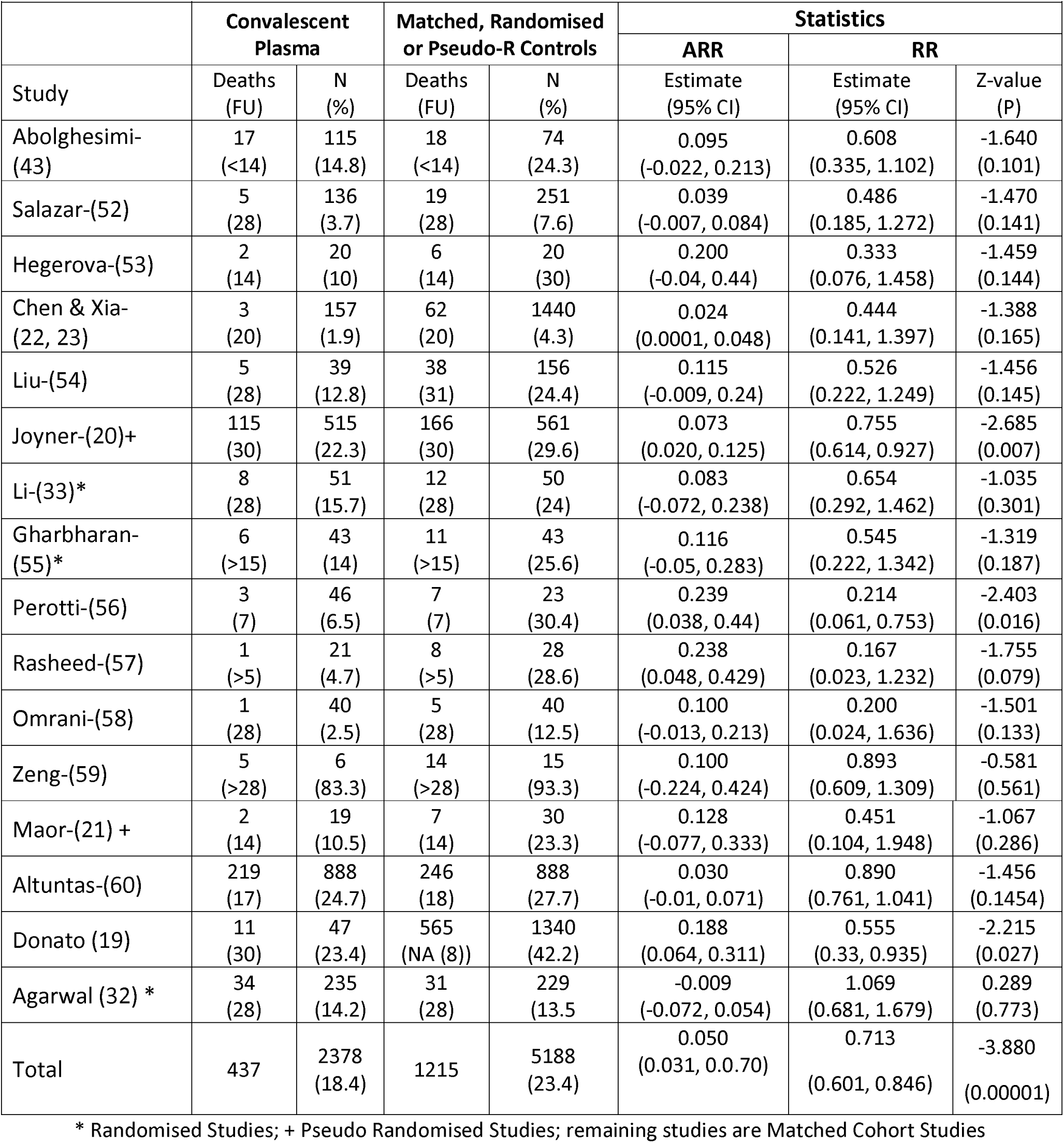
Meta-analysis Convalescent Plasma vs Matched/randomized*/pseudo-randomised+ series

The most statistically significant study was the first pseudo-randomised study with 515 high titre vs 561 low dose titre plasma (Table 1, 22.3% vs 29.6% deaths after 30 days follow up with RR = 0.755 (95% CI: 0.614, 0.927), z-value = −3.528 and p= 0.007). This was even larger when the difference in time to transfusion, =<3 days verses > 4days of COVID-19 diagnosis was also taken into account in a further “pseudo-pseudo” randomisation (Table 2 16.7% in 180 patients vs 31.8% death rate in 371 patients at 30 days RR = 0.524 (95% CI: 0.366, 0.750) p= 0.0004).

**Table 2.** Subset analysis of interaction of =<3 verses 4+ days delay in Convalescent plasma therapy and high versus low (HD vs LD) titre plasma on 30-day mortality (20)

|  |  |  |  |  |
| --- | --- | --- | --- | --- |
|  | HD Convalescent plasma death rate (n/N) | LD Convalescent death rate (n/N) | Absolute risk reduction | RR (95% CI) |
| <= 3days | 16.7% (30/180) | 25.3% (48/190) | 8.4% | 0.660<br>(0.439, 0.992) |
| 4+ days | 25.4% (85/335) | 31.8% (118/371) | 6.4% | 0.798<br>(0.630, 1.011) |
| <=3 days high vs 4+ days low* | 16.7% (30/180) | 31.8% (118/371) | 15.1% | 0.524<br>(0.366, 0.750) |
| **"pseudo-pseudo" randomisation (see appendix 1 ) |  |  |  |  |

Despite only one other study being statistically significant individually, all of the smaller studies had approximately a similar percentage risk reduction (Table 1).

We performed four random meta-analyses using the restricted maximum likelihood method estimator. The pooled estimate of all study types (i.e. matched cohort studies, randomised trials and pseudo randomised trials) shows a statistically significant reduction in COVID-19 deaths with a RR of 0.713 (95% CI: 0.601, 0.846), z-value = −3.880 and p-value = 0.0001. Similarly, a statistically significant reduction in COVID-19 deaths from the eleven matched cohort studies meta-analysis with a RR of 0.610 (95% CI: 0.459, 0.810), z-value = −3. 418 and p-values = 0.001. However, the result of the meta-analysis of the three randomised trials did not show a statistically significant reduction of COVID-19 deaths RR = 0.825 (95% CI: 0.529, 1.287), z-value = −0.846 and p-value = 0.397. When the two pseudo-randomised studies were pooled together, there were a statistically significant reduction in COVID-19 deaths RR = 0.747 (95% CI: 0.610, 0.916), z-value = −2.807 and p-value = 0.005.

The result of the meta-regression mixed-effects model estimated no statistically significant differences between the three study types. The estimated effect of the pseudo-randomised studies compared with the matched cohort studies was 0.141 (95% CI: −0.442, 0.724) with z-value = 0.473 and p-value = 0.636. Similarly, no statistically significant difference was observed between the randomised trials and the reference group the matched cohort studies 0.267 (95% CI: −0.291, 0.825), z-value = 0. 938 and p-value = 0.348. The estimated I^2^ = 40.40% indicating non-statistically significant moderate observed heterogeneity with p-value = 0.092 of the test statistic, which examines the null hypothesis that all studies are evaluating the same effect.

A comparison of the mortality rate from convalescent plasma studies and the currently licenced products Dexamethasone and Remdesivir is shown in Table 3 and a comparison of recovery times of convalescent plasma vs Remdesivir and the monoclonal antibody Regeneron is shown in Table 4

**Table 3:**
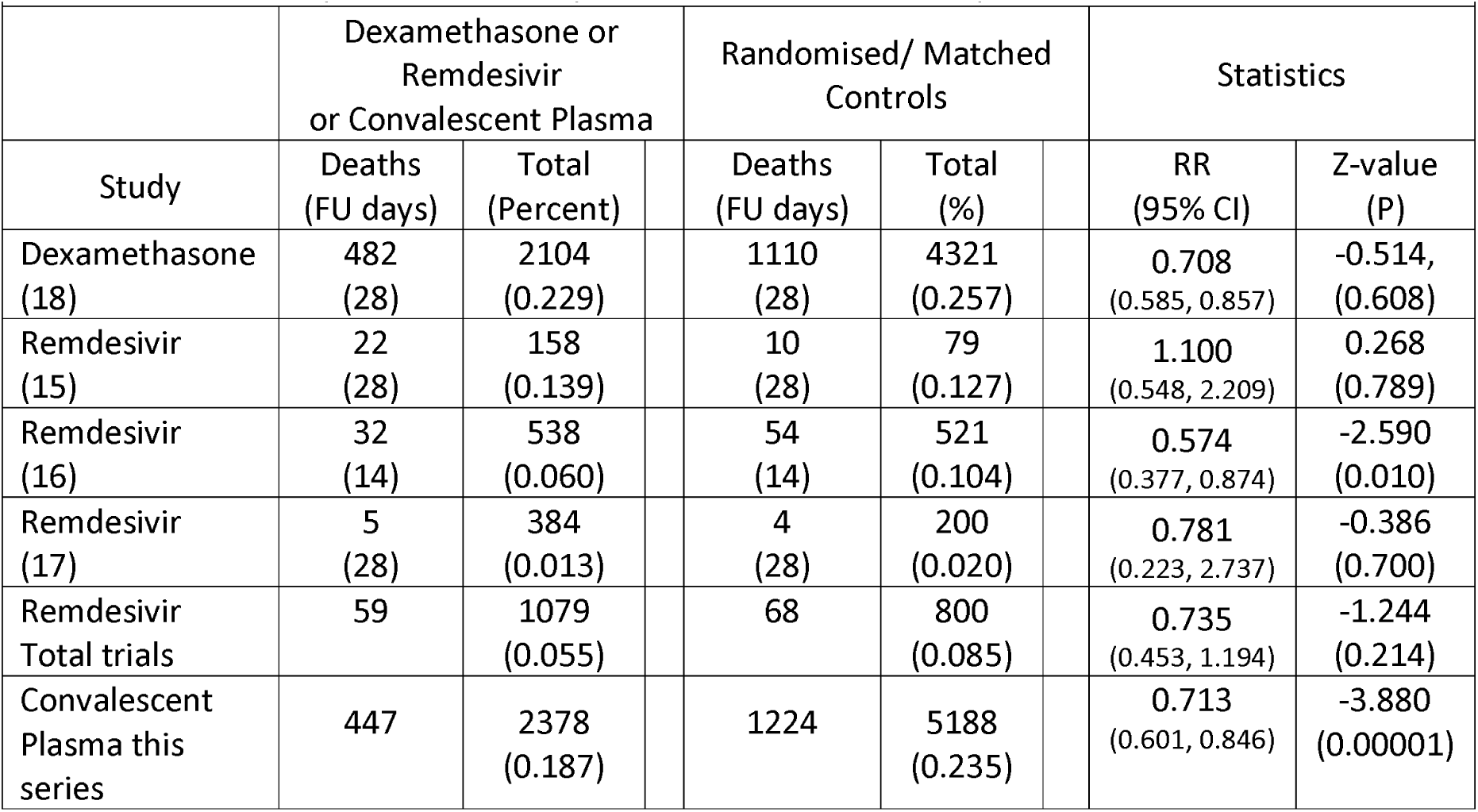
Comparison of mortality after Convalescent Plasma/Dexamethasone/Remdesivir vs Randomized/Matched controls

**Table 4.** Comparative recovery times versus controls after Monoclonal antibodies, Convalescent Plasma and Remdesivir

|  |  |  |  |  |  |  |  |
| --- | --- | --- | --- | --- | --- | --- | --- |
|  | Time to recovery treatment | Total No | Percent improvement | Time to recover controls | Total No | Statistic | P value |
| Monoclonal antibody * | 7 days | NA | 0.461 | 13 days | NA | NA | NA |
| Convalescent Plasma (43) | 6.25<sup>+</sup> days | 115 | 0.515<sup>+</sup> | 12.88 days | 74 | t-test | P<0.0006<sup>+</sup> |
| Remdesivir (16) | 11 days | 538 | 0.267 | 15 days | 521 | HR 1.32 (1.23, 1.55) | P < 0.001 |
| [*http://www.prnewswire.com/news-releases/regenerons-regn-cov2-antibody-cocktail-reduced-viral-levels-and-improved-symptoms-in-non-hospitalized-covid-19-patients-301140336.html](http://www.prnewswire.com/news-releases/regenerons-regn-cov2-antibody-cocktail-reduced-viral-levels-and-improved-symptoms-in-non-hospitalized-covid-19-patients-301140336.html)<br><sup>+</sup> time from Convalescent Plasma transfusion to discharge |  |  |  |  |  |  |  |

Two studies investigated the impact of Convalescent Plasma on persistence of COVID-19 Viral nucleic acid detection. Both showed faster decline compared to control but only one showed a statistically significant effect at 72 hours. (Table 5).

**Table 5:** Impact of Convalescent Plasma (CP) on clearance Virus nucleic acid (32, 33)

|  |  |  |  |  |  |  |
| --- | --- | --- | --- | --- | --- | --- |
| Li et al (33) | All cases |  | Severe Disease |  | Life-threatening disease |  |
| Viral negative | Convalescent Plasma (n=47) | Control Group (n=40) | Convalescent Plasma (n=21) | Control Group (n=17) | CP (n=26) | Control Group (n=23) |
| 24 hrs | 44.7% | 15% | 33.3% | 11.8% | 53.8% | 17.4% |
| 48 hrs | 68.1% | 32.5% | 61.9% | 35.3% | 73.1% | 30.4% |
| 72hrs | 87.2% | 37.5% | 90.5% | 41.2% | 84.6% | 34.8% |
| Stats | Odds Ratio (95% CI), p-value |  | Odds Ratio (95% CI), p-value |  | OR (95% CI), p-value |  |
| 24 hrs | 4.58 (1.62-12.96), p=0.003 |  | 3.75 (0.66-21.20), 0.150 |  | 5.54 (1.47-20.86), 0.010 |  |
| 48 hrs | 4.43 (1.80-10.92), p=0.001 |  | 2.98 (0.79-11.25), 0.100 |  | 6.20 (1.79-21.46), 0.003 |  |
| 72 hrs | 11.39 (3.91-33.18), p<0.001 |  | 13.57 (2.36-77.95), <0.001 |  | 10.31(2.63-40.5), <0.001 |  |
| Agarwal (32) | Convalescent Plasma (n=184) | Control Group (n=183) |  |  |  |  |
| 72 hrs | 42.9% | 36.6% |  |  |  |  |
| Stats | Odds Ratio (95% CI), p-value |  |  |  |  |  |
| 72 hrs | 1.303 (0.857, 1.981), p= 0.216 |  |  |  |  |  |

## Discussion

With more than a million COVID-19 deaths world wide (https://www.ecdc.europa.eu/en/geographical-distribution-2019-ncov-cases) and more than 6,000 a day, there is a need for more than a vaccine to control this pandemic. While agreeing with recent editorials calling for more effort to recruit to randomised trials with Convalescent Plasma (9, 31), it is clear from the figures in Table 1 (only 329 of 2378 – 13.8% receiving convalescent plasma in a randomised trial) that, faced with the initial high mortality rate, most clinicians actually treating patients preferred first to demonstrate benefit compared to untreated but matched controls. By no means, without results from larger series of randomised trials, can the mixed standards of reporting in this systematic review be trusted on their own. Some studies had unknown antibody titre, there was differences in length of follow-up and confounding by co-administration of other COVID-19 treatments in the Matched cohort series. They are certainly all biased to a degree as indicated by those included in the Cochrane review which included series up to the beginning of August (11). However, in the absence of any better material, accepting these imperfections, from the meta-analyses of death rates there is a highly significant reduction of death in the period up to 30 days after transfusion (Table 3 RR= 0.713,, Z= −3.680 p< 0.0001) Furthermore, this is not far from the non-significant findings reported in two of the under-powered randomised studies (RR 0.654 and 0.545) though it is better than the Indian randomised trial reported by Agarwal et al Table 1 (RR 1.069 (32)). This trial used young donors who only had mild symptoms and 72% of recipients received plasma with a low titre or no antibody when checked retrospectively.

Despite the significant reduction of deaths in the pooled analysis, the reduction in death is less than the odds ratio, 0.25; 95% confidence interval, 0.14–0.45; I^2^ = 0% p< 0.001 reported from matched cohort studies of convalescent plasma before the COVID-19 pandemic (4). However, like these studies, the COVID-19 randomised data from Li et al (33) and “pseudo-randomised” data from Joyner et al (20) do suggest greater benefit from treating less extreme cases, treating within 3 days of onset of illness and use of high antibody titre sera. The current results are certainly better when compared to those reported in both the dexamethasone trial and the Remdesivir trials (15, 17, 34) (Table 3). This is most critically emphasised in the study of Wang et al (15) which showed absolutely no difference in the rate of decline of Viral load following Remdesivir than in the control group over 28 days. As table 5 demonstrates Li et al (33) demonstrated a markedly different effect from the study of convalescent plasma when significant reductions are apparent within 24 hours and 87.2% of CP treated compared to 31.8% in the randomised control group being negative within 72 hours (33). This advantage was present both in early severe disease and advanced life-threatening disease reaching p<.001 in both groups by 72 hours. Despite this similar early effect, the latter group had slower recovery and did not show survival advantage at 30 days follow up possibly because of more systemic damage. The study of viral disappearance was markedly less impressive in Agarwal et al’s trial (32). This was the only trial showing no reduction of mortality. A possible explanation could have been the low rate of high titre anti-COVID-19 antibody in their donors (26.7% had no detectable antibody and only 28.0% had titres >1:80)

Critical to the justification of pursuing randomised trials is the need to confirm that the complications of convalescent plasma are as low as reported (11, 35) particularly given the anxiety from animal studies about immune enhancement (36) and serum screening of COVID-19 survivors have detected autoantibodies to Interferon (37). This will be important to rule out, given that Convalescent plasma is much less toxic than those seen in the randomised trials supporting the two licenced drugs. These showed possible (though non-significant) harm to patients not receiving respiratory support while undergoing Dexamethasone (18) and 21% of patients in the 5-day group and 35% in the 10-day group having serious adverse events while undergoing Remdesivir treatment (38).

Hopefully, this report will help accelerate completion of more than 4 randomised trials involving nearly 4,000 patients. These were expected within the next 1-3 months but recruitment slowed due to a Summer decline in advanced cases in the northern hemisphere (39) and the need to select only those donors with high titre neutralising antibodies against the SARS-CoV-2 spike protein. It is hoped that these can be completed soon as trials with the high titre monoclonal antibodies (40) are now beginning (41, 42). Though no data has been published in a scientific journal, a press release of early results from treating COVID-19 antibody-negative non-hospitalised patients are indicating activity (https://investor.regeneron.com/news-releases/news-release-details/regenerons-regn-cov2-antibody-cocktail-reduced-viral-levels-and/). The only possible direct comparison between these early results and this report is when the results of time to recovery are examined. After the monoclonal antibody treatment versus untreated controls there is 7 vs 13 days, 46.2% reduction. This compares to 11 vs 15 days (26.7% reduction) after Remdesivir (34) and 6.25 vs 12.88 days, (51.5% reduction in a CP trial (43) (Table 4). Completing the Convalescent plasma trials will also be important for future epidemics as convalescent sera can become available more quickly than monoclonal antibodies. This is particularly important for areas without access to western-style medicine (44). Equally, there could be something in recovered plasma which may be related to other determinants on the virus that contributes to virulence than the monoclonal antibody recognises. This means that it will be important that monoclonals are tested against CP in randomised trials. The most sensitive way of testing these two approaches could be a 2×2 randomised trial of high titre CP versus Monoclonal and a second randomisation for immediate versus deferred use based on the rate of decline in Virus titre over 72 hours.

Missing information from the currently available published data is the impact of convalescent plasma in patients with extreme poor risk factors such as obesity, diabetes, old age and ethnicity. In the US EAP COVID-19 consortium study, there is double the death rate in over 70s though just under 2/3rds were alive after 30 days verses 4/5^th^ of the under 70s (20). In addition, there has so far been no study providing long term survival and quality of life data in CP patients, given the increasing recognition of the problem of late persistent poor quality of life in a significant minority of recovered patients (45).

The final issue worth further investigation is the impact of Vitamin D deficiency, as there have been no studies of the influence of Vitamin deficiency on failure to respond to Convalescent plasma. In the last month, there have been increasing interest (46) with six reports demonstrating a strong impact of Vitamin D deficiency on the behaviour of SARS-CoV-2 infection. Firstly, Meltzer et al studied Vitamin D levels in 489 attendees in an urban medical centre in Chicago (47). They demonstrated a higher frequency of acquisition of COVID-19 acquisition in the Vitamin D deficient group (21.9%) than in the sufficient group (12.2%). This was subsequently confirmed by a much larger study (48) and two smaller studies (46, 49). The Boston group also reported data suggesting a link between Vitamin D deficiency and increased severity of disease (50). This might explain the failure to demonstrate any short term survival benefit of convalescent plasma in patients with life-threatening disease in the randomised trial of Li et al despite equal efficiency in dealing with actual presence of the virus within 72 hours of transfusion (Table 5). It might be important if this prediction proves to be true, as patients with life-threatening disease could respond better to treatment with more rapidly acting Calcifediol rather than Vitamin D3 given the recent report from Spain (51). These authors reported a small randomised study which demonstrated that only 1 of 50 hospitalised COVID-19 patients receiving Colifediol required admission to intensive care unit versus 13 of 26 untreated controls (p< 0.001)

## Conclusion

It is 10 months since this pandemic and worldwide there continue to be close to 6,000 deaths a day and total of more than 1 million deaths

The data analysed in this paper suggests CP given within 3 days of admission at an adequate titre could reduce deaths by as much as 40% and is better than that achieved from any other licenced drug, with less side effects

Expansion of current randomised trials and more education amongst potential donors and the medical profession in general to enable more widespread access to such trials are required as we enter the second wave of this pandemic.

## Data Availability

contact

## Authors Contribution

each author contributed equally, MS & TO primarily extracting data and AA undertaking statistical analysis

## References

1. Behring Ev, Kitasato S. On the Development of Diphtheria Immunity and Tetanus Immunity in Animals. Deutsche Medizinische Wochenschrift 1890(49):1–7.

2. Cavaillon JM. Historical links between toxinology and immunology. Pathogens and Disease. 2018;76(3):1–12.

3. Flexner S. The Results of the Serum Treatment in Thirteen Hundred Cases of Epidemic Meningitis. J Exp Med. 1913;17(5):553–76.

4. Mair-Jenkins J, Saavedra-Campos M, Baillie JK, Cleary P, Khaw FM, Lim WS, et al. The effectiveness of convalescent plasma and hyperimmune immunoglobulin for the treatment of severe acute respiratory infections of viral etiology: a systematic review and exploratory meta-analysis. J Infect Dis. 2015;211(1):80–90.

5. van Griensven J, Edwards T, de Lamballerie X, Semple MG, Gallian P, Baize S, et al. Evaluation of Convalescent Plasma for Ebola Virus Disease in Guinea. N Engl J Med. 2016;374(1):33–42.

6. Luke TC, Kilbane EM, Jackson JL, Hoffman SL. Meta-analysis: convalescent blood products for Spanish influenza pneumonia: a future H5N1 treatment? Ann Intern Med. 2006;145(8):599–609.

7. Duan K, Liu B, Li C, Zhang H, Yu T, Qu J, et al. Effectiveness of convalescent plasma therapy in severe COVID-19 patients. Proc Natl Acad Sci U S A. 2020.

8. Joyner MJ, Klassen SA, Johnson PWea. Evidence favouring the efficacy of convalescent plasma for COVID-19 therapy. medRxiv [Internet]. 2020:p[1-9 pp.].

9. Roberts DJ, Miflin G, Estcourt L. Convalescent plasma for COVID-19: Back to the future. Transfus Med. 2020;30(3):174–6.

10. Valk SJ, Piechotta V, Chai KL, Doree C, Monsef I, Wood EM, et al. Convalescent plasma or hyperimmune immunoglobulin for people with COVID-19: a rapid review. Cochrane Database Syst Rev. 2020;5:CD013600.

11. Piechotta V, Chai KL, Valk SJ, Doree C, Monsef I, Wood EM, et al. Convalescent plasma or hyperimmune immunoglobulin for people with COVID-19: a living systematic review. Cochrane Database Syst Rev. 2020;7:CD013600.

12. Chai KL, Valk SJ, Piechotta V, Kimber C, Monsef I, Doree C, et al. Convalescent plasma or hyperimmune immunoglobulin for people with COVID-19: a living systematic review. Cochrane Database Syst Rev. 2020;10:CD013600.

13. Pal P, Ibrahim M, Niu A, Zwezdaryk KJ, Tatje E, Robinson WRt, et al. Safety and efficacy of COVID-19 convalescent plasma in severe pulmonary disease: A report of 17 patients. Transfus Med. 2020.

14. Focosi D, Farrugia A. ABO-incompatible convalescent plasma transfusion: Yes, you can. Transfus Med. 2020.

15. Wang Y, Zhang D, Du G, Du R, Zhao J, Jin Y, et al. Remdesivir in adults with severe COVID-19: a randomised, double-blind, placebo-controlled, multicentre trial. Lancet. 2020;395(10236):1569–78.

16. Beigel JH, Tomashek KM, Dodd LE, Mehta AK, Zingman BS, Kalil AC, et al. Remdesivir for the Treatment of Covid-19 - Final Report. N Engl J Med. 2020.

17. Spinner CD, Gottlieb RL, Criner GJ, Arribas Lopez JR, Cattelan AM, Soriano Viladomiu A, et al. Effect of Remdesivir vs Standard Care on Clinical Status at 11 Days in Patients With Moderate COVID-19: A Randomized Clinical Trial. JAMA. 2020;324(11):1048–57.

18. Group RC, Horby P, Lim WS, Emberson JR, Mafham M, Bell JL, et al. Dexamethasone in Hospitalized Patients with Covid-19 - Preliminary Report. N Engl J Med. 2020.

19. Donato M, Park S, Baker M, al. e. Clinical and laboratory evaluation of patients with SARS-CoV-2 pneumonia treated with high-titer convalescent plasma: a prospective study. meDxiv [Internet]. 2020:p[1-13 pp.].

20. Joyner MJ, Senefeld JW, Klassen SA, Mills JR, Johnson PW, Theel ES, et al. Effect of Convalescent Plasma on Mortality among Hospitalized Patients with COVID-19: Initial Three-Month Experience. medRxiv. 2020.

21. Maor Y, Cohen D, Paran N, Israely T, Ezra V, Axelrod O, et al. Compassionate use of convalescent plasma for treatment of moderate and severe pneumonia in COVID-19 patients and association with IgG antibody levels in donated plasma. EClinicalMedicine. 2020;26:100525.

22. Chen B, Xia R. Early experience with convalescent plasma as immunotherapy for COVID-19 in China: Knowns and unknowns. Vox sanguinis. 2020.

23. Xia X, Li K, Wu L, Wang Z, Zhu M, Huang B, et al. Improved clinical symptoms and mortality among patients with severe or critical COVID-19 after convalescent plasma transfusion. Blood. 2020;136(6):755–9.

24. Berkey CS, Hoaglin DC, Mosteller F, Colditz GA. A random-effects regression model for meta-analysis. Statistics in medicine. 1995;14(4):395–411.

25. Altman DG. Practical statistics for medical research.: Chapman and Hall London; 1991.

26. Grant RL. Converting an odds ratio to a range of plausible relative risks for better communication of research findings. BMJ 2014: f7450 [Internet]. 2014.

27. Higgins JP, Thompson SG, Deeks JJ, Altman DG. Measuring inconsistency in meta-analyses. BMJ. 2003;327(7414):557–60.

28. Borenstein M, Hedges LV, Higgins JP, R. RH. Introduction to Meta-Analysis: John Wiley & Sons.; 2011.

29. RCT. R: A language and environment for statistical computing. R Foundation for Statistical Computing, Vienna, Austria. 2020.

30. Chen B, Xia R. Early experience with convalescent plasma as immunotherapy for COVID-19 in China: Knowns and unknowns. Vox Sang. 2020;115(6):507–14.

31. Estcourt LJ, Roberts DJ. Convalescent plasma for covid-19. BMJ. 2020;370:m3516.

32. Agarwal A, Mukherjee A, Kumar Gea. Convalescent plasma in the management of moderate covid-19 in adults in India: open label phase II multicentre randomised controlled trial (PLACID Trial). BMJ. 2020;371:m3939.

33. Li L, Zhang W, Hu Y, Tong X, Zheng S, Yang J, et al. Effect of Convalescent Plasma Therapy on Time to Clinical Improvement in Patients With Severe and Life-threatening COVID-19: A Randomized Clinical Trial. JAMA. 2020;324(5):460–70.

34. Beigel JH, Tomashek KM, Dodd LE, Mehta AK, Zingman BS, Kalil AC, et al. Remdesivir for the Treatment of Covid-19 - Preliminary Report. N Engl J Med. 2020.

35. Joyner MJ, Bruno KA, Klassen SA, Kunze KL, Johnson PW, Lesser ER, et al. Safety Update: COVID-19 Convalescent Plasma in 20,000 Hospitalized Patients. Mayo Clin Proc. 2020;95(9):1888–97.

36. Arvin AM, Fink K, Schmid MA, Cathcart A, Spreafico R, Havenar-Daughton C, et al. A perspective on potential antibody-dependent enhancement of SARS-CoV-2. Nature. 2020;584(7821):353–63.

37. Bastard P, Rosen LB, Zhang Q, Michailidis E, Hoffmann HH, Zhang Y, et al. Auto-antibodies against type I IFNs in patients with life-threatening COVID-19. Science. 2020.

38. Goldman JD, Lye DCB, Hui DS, Marks KM, Bruno R, Montejano R, et al. Remdesivir for 5 or 10 Days in Patients with Severe Covid-19. N Engl J Med. 2020.

39. Cunniffe NG, Gunter SJ, Brown M, Burge SW, Coyle C, De Soyza A, et al. How achievable are COVID-19 clinical trial recruitment targets? A UK observational cohort study and trials registry analysis. BMJ Open. 2020;10(10):e044566.

40. Zost SJ, Gilchuk P, Chen RE, Case JB, Reidy JX, Trivette A, et al. Rapid isolation and profiling of a diverse panel of human monoclonal antibodies targeting the SARS-CoV-2 spike protein. Nat Med. 2020;26(9):1422–7.

41. Sewell HF, Agius RM, Kendrick D, Stewart M. Vaccines, convalescent plasma, and monoclonal antibodies for covid-19. BMJ. 2020;370:m2722.

42. Marovich M, Mascola JR, Cohen MS. Monoclonal Antibodies for Prevention and Treatment of COVID-19. JAMA. 2020;324(2):131–2.

43. Abolghasemi H, Eshghi P, Cheraghali AM, Imani Fooladi AA, Bolouki Moghaddam F, Imanizadeh S, et al. Clinical efficacy of convalescent plasma for treatment of COVID-19 infections: Results of a multicenter clinical study. Transfusion and apheresis science: official journal of the World Apheresis Association: official journal of the European Society for Haemapheresis. 2020:102875.

44. Hassan MO, Osman AA, Elbasit HEA, Hassan HE, Rufai H, Satti MMM, et al. Convalescent plasma as a treatment modality for coronavirus disease 2019 in Sudan. Transfus Apher Sci. 2020:102918.

45. Arnold DT, Hamilton F.W, Milne Aea. Patient outcomes after hospitalisation with COVID-19 and implications for follow-up; results from a prospective UK cohort. medRxiv 2020.

46. Panagiotou G, Tee SA, Ihsan Y, Athar W, Marchitelli G, Kelly D, et al. Low serum 25-hydroxyvitamin D (25[OH]D) levels in patients hospitalized with COVID-19 are associated with greater disease severity. Clin Endocrinol (Oxf). 2020.

47. Meltzer DO, Best TJ, Zhang H, Vokes T, Arora V, Solway J. Association of Vitamin D Status and Other Clinical Characteristics With COVID-19 Test Results. JAMA Netw Open. 2020;3(9):e2019722.

48. Kaufman HW, Niles JK, Kroll MH, Bi C, Holick MF. SARS-CoV-2 positivity rates associated with circulating 25-hydroxyvitamin D levels. PLoS One. 2020;15(9):e0239252.

49. Merzon E, Tworowski D, Gorohovski A, Vinker S, Golan Cohen A, Green I, et al. Low plasma 25(OH) vitamin D level is associated with increased risk of COVID-19 infection: an Israeli population-based study. FEBS J. 2020.

50. Maghbooli Z, Sahraian MA, Ebrahimi M, Pazoki M, Kafan S, Tabriz HM, et al. Vitamin D sufficiency, a serum 25-hydroxyvitamin D at least 30 ng/mL reduced risk for adverse clinical outcomes in patients with COVID-19 infection. PLoS One. 2020;15(9):e0239799.

51. Entrenas Castillo M, Entrenas Costa LM, Vaquero Barrios JM, Alcala Diaz JF, Lopez Miranda J, Bouillon R, et al. “Effect of calcifediol treatment and best available therapy versus best available therapy on intensive care unit admission and mortality among patients hospitalized for COVID-19: A pilot randomized clinical study”. J Steroid Biochem Mol Biol. 2020;203:105751.

52. Salazar E, Christensen PA, Graviss EA, Nguyen DT, Castillo B, Chen J, et al. Treatment of Coronavirus Disease 2019 Patients with Convalescent Plasma Reveals a Signal of Significantly Decreased Mortality. Am J Pathol. 2020.

53. Hegerova L, Gooley TA, Sweerus KA, Maree C, Bailey N, Bailey M, et al. Use of convalescent plasma in hospitalized patients with COVID-19: case series. Blood. 2020;136(6):759–62.

54. Liu STH, Lin HM, Baine I, Wajnberg A, Gumprecht JP, Rahman F, et al. Convalescent plasma treatment of severe COVID-19: a propensity score-matched control study. Nat Med. 2020.

55. Gharbharan Hea. Convalescent Plasma for COVID-19. medRxiv. 2020:1–16.

56. Perotti C, Baldanti F, Bruno R, Del Fante C, Seminari E, Casari S, et al. Mortality reduction in 46 severe Covid-19 patients treated with hyperimmune plasma. A proof of concept single arm multicenter trial. Haematologica. 2020.

57. Rasheed AM, Fatak DF, Hashim HA, Maulood MF, Kabah KK, Almusawi YA, et al. The therapeutic potential of convalescent plasma therapy on treating critically-ill COVID-19 patients residing in respiratory care units in hospitals in Baghdad, Iraq. Le infezioni in medicina: rivista periodica di eziologia, epidemiologia, diagnostica, clinica e terapia delle patologie infettive. 2020;28(3):357–66.

58. Omrani AS, Zaqout A, Baiou A, Daghfal J, Elkum N, Alattar RA, et al. Convalescent plasma for the treatment of patients with severe coronavirus disease 2019: A preliminary report. J Med Virol. 2020.

59. Zeng QL, Yu ZJ, Gou JJ, Li GM, Ma SH, Zhang GF, et al. Effect of Convalescent Plasma Therapy on Viral Shedding and Survival in Patients With Coronavirus Disease 2019. J Infect Dis. 2020;222(1):38–43.

60. Altuntas F, Ata N, Yigenoglu TN, Basci S, Dal MS, Korkmaz S, et al. Convalescent plasma therapy in patients with COVID-19. Transfus Apher Sci. 2020:102955. (https://www.ncbi.nlm.nih.gov/pmc/articles/PMC7456194/)

